# Countrywide quarantine only mildly increased anxiety level during COVID-19 outbreak in China

**DOI:** 10.1101/2020.04.01.20041186

**Authors:** Wei Hu, Li Su, Juan Qiao, Jing Zhu, Yi Zhou

## Abstract

In the recent outbreak of COVID-19, many countries have taken various kinds of quarantine measures to slow down the explosive spreading of COVID-19. Although these measures were proven to be successful in stopping the outbreak in China, the potential adverse effects of countrywide quarantine have not been thoroughly investigated. In this study, we performed an online survey to evaluate the psychological effects of quarantine in China using Zung Self-rating Anxiety Scale in February 2020 when the outbreak was nearly peaked in China. Along with the anxiety scores, limited personal information such as age, gender, region, education, occupation and specifically, the type and duration of quarantine were collected for analysis. For a total number of 992 valid questionnaires, clinical significance of anxiety symptoms was observed in 9.58% respondents according to clinical diagnostic standards in China. Statistical results showed population with different age, education level, health status and personnel category responded differently. Other characteristics such as gender, marital status, region, and acquaintance with suspected or confirmed cases of COVID-19 did not affect anxiety levels significantly. Respondents experienced different forms of quarantine showed different anxiety levels. Unexpectedly, longer durations of quarantine did not lead to significant increase of anxiety level. Our results suggest a rather mild psychological influence caused by the countrywide quarantine during COVID-19 outbreak in China and provided reference for other countries and regions to battle COVID-19.

## Introduction

On March 11^th^ 2020, the World Health Organization declared the SARS-COV-2 induced COVID-19 as a “pandemic” during a news conference in Geneva(European Centre for Disease Prevention and Control, 2020). With more than 120,000 confirmed infections and more than 5,000 lives taken in more than 100 countries, there is still no specific medicine to cure the highly transmissive COVID-19 (Zhang and Liu, 2020). Although some promising drugs (e.g. Remdesivir, Favipiravir) are in clinical trials now (Wang et al., 2020, Dong et al., 2020), the most effective way to stop COVID-19 by far, is still the oldest way that human being had used to battle epidemics for hundreds of years: quarantine. China has been conducting quarantine in many provinces over the whole country since late January and the results are significant. In about two weeks, daily number of new patients peaked and then began to decline (World Health Organization, 2020). Six weeks after quarantine, daily number of new patients has been dropped to less than 100 in China (National Health Commission of the People’s Republic of China, 2020a). Similar quarantine methods have also been adopted by other countries like Republic of Korea. Since the beginning of regional quarantine in Daegu, the number of newly diagnosed cases in Republic of Korea has been declining steadily as well(Choi and Ki, 2020). Other countries like Iran (countrywide quarantine since March, 2020) and Italy (countrywide quarantine since March, 2020) are also taking quarantine measures recently and more countries might also take these approaches into consideration to stop COVID-19 outbreak.

However, it was never an easy option to make a decision to quarantine due to predictable huge negative impacts to economy and unpredictable psychological harm to quarantined population. Previous reports have shown that negative emotions caused by quarantine can lead to various kinds of consequences such as anxiety, depression and posttraumatic stress disorder (PTSD)(Brooks et al., 2020). During the outbreak of SARS (severe acute respiratory syndrome) in 2003, an increased prevalence of depression and PTSD was found in quarantined persons(Hawryluck et al., 2004). During MERS (middle east respiratory syndrome) outbreak, quarantined people showed more negative emotions such as anxiety and anger(Jeong et al., 2016). Evidence from animal experiments even showed a 30% decrease of neuron cells in the brain in isolated mouse (Heng et al., 2018). These evidences suggested that potential psychological and other effects of quarantine could be significant and should be carefully taken into consideration. However, very different from the quarantine measures during SARS or MERS outbreak, a much larger population in China are affected by the countrywide quarantine to battle COVID-19. A rough estimation is that more than 100 million people in China were affected by this countrywide quarantine(Tian et al., 2020) which no one has ever seen in human history. This raises new concerns regarding the potential adverse effects of large-scale quarantine. Will countrywide quarantine cause broad social panic and even chaos? Or oppositely, super large-scale quarantine might have no major impacts to general public because everyone was quarantined — according to the equity theory (Adams, 1965). These controversial questions cannot be easily answered without systematic investigation but could be critical important for future decision of large-scale quarantine.

In this study, we performed an online questionnaire survey during the middle stage of COVID-19 outbreak in China (Fig. S1) to understand the psychological effect on quarantined persons using Zung Self-rating Anxiety Scale (Zung, 1971). Along with the anxiety scores, limited personal information such as age, gender, region, education etc. and specifically, the type and durations of quarantine were collected for analysis. We further assessed the influence of different types and different durations of quarantine used in China. Our results provided new insights about understanding experiences of quarantined persons during COVID-19 outbreak in China which could be important for global-wide containment of COVID-19 in other countries.

## Method

### Survey tool

The survey was conducted in the form of an online questionnaire between 12:00PM February 19^th^ 2020 and 12:00PM February 26^th^ 2020 (UTC+8), which consisted of a general information survey and a Zung Self-Rating Anxiety Scale (SAS) (Zung, 1971). The general survey includes: 1. basic information of the respondent, such as gender, age, region, education level, occupation, marital status and phone number (optional) for more contact. 2. Quarantine related information of the respondent during COVID-19 outbreak. Anxiety assessment was performed using the Self-Rating Anxiety Scale (SAS) compiled by William W. K. Zung. SAS is used to measure the subjective anxiety of subjects, using a 4-point scale: no or very little time, a small amount of time, a considerable amount of time, most or all of the time. Raw scores were then converted to index scores following previous report (Zung, 1971). Index score equal or larger than 50 is considered as clinical significance of anxiety (Zung, 1986), 50-59 is mild anxiety, 60-69 is moderate anxiety, and 70 or more is severe anxiety.

### Investigation methods

The online survey was performed using a professional online survey service Questionnaire Star (https://www.wjx.cn), and then released nationwide through social software (such as WeChat, Weibo, QQ, etc.). This study was conducted with informed consent of the respondent.

### Quality Control

IP address was often used for quality control in online survey. Considering that quarantined personnel or families might share the same internet and same IP address, we did not limit the number of questionnaires from the same IP address. Instead, we performed post hoc check to ensure the reliability of questionnaires. For a total number of 997 questionnaires collected, 5 were identified as invalid questionnaires due to abnormal key data (e.g. 4 years old). For a total number of 888 unique IP addresses, 816 (82.26%) IP addresses filed 1 questionnaire, 55 (6.19%) filed 2, 10 (1.13%) filed 3, 7 (0.79%) filed 4 or more (max 7). The median time to finish this questionnaire was 291 seconds with interquartile range from 215 – 415 seconds.

### Statistical methods

Statistical analysis was performed using SPSS 22 (IBM, USA). Cronbach’s alpha was used to measure the reliability. The KMO (Kaiser-Meyer-Olkin) value and Bartlett’s sphericity test were used to examine the suitability of data. Independent sample t-test was use for analysis of gender, region, health status, and acquaintance with suspected or confirmed cases of COVID-19. For multiple groups of samples (age, education, marital status, and personnel category), one-way ANOVA analysis was used. The confidence level was set at P < 0.05 unless specified.

## Result

### 1. Demographics and description of respondents

The questionnaire survey collected 992 valid questionnaires (see Methods for details) between 12:00PM February 19^th^ 2020 and 12:00PM February 26^th^ 2020 (UTC+8). Cronbach’s alpha is equal to 0.812 suggesting the reliability is robust. The KMO value was 0.917 indicate the sampling is adequate. Bartlett’s sphericity test was considered statistically significant with p value smaller than 0.001.

Table 1 summarized personal characteristics from valid questionnaires. There are 424 males and 568 females. The age ranged from 11 to 75 years, with a median age of 36 years (IQR: interquartile range, 28 - 42). There are 214 (21.6%) from Hubei Province, the province with the most severe outbreak (>60,000 infections), and 778 (78.4%) from other regions in China. The education levels of 75 (7.6%) respondents are junior high school or below, 177 (17.8%) are high school / technical secondary school, 247 (24.9%) are college / higher vocational, 423 (42.6%) are undergraduate, 70 (7.1%) are postgraduate. In terms of marital status, 700 were married (70.6%), 252 were unmarried (25.4%), 37 were divorced (3.7%), and 3 were widowed (0.3%). For personnel category, 41 (4.1%) and 102 (10.3%) were frontline medical and non-medical personnel for battling COVID-19 respectively, 106 (10.7%) were non-frontline medical personnel, and 743 (74.9%) were others. In terms of healthy status, 961 (96.9%) are healthy, and 31 (3.1%) had chronic diseases. Among all respondents, 81 (8.2%) have acquaintance with people diagnosed or suspected with COVID-19, and 911 (91.8%) were not.

**Table 1.**
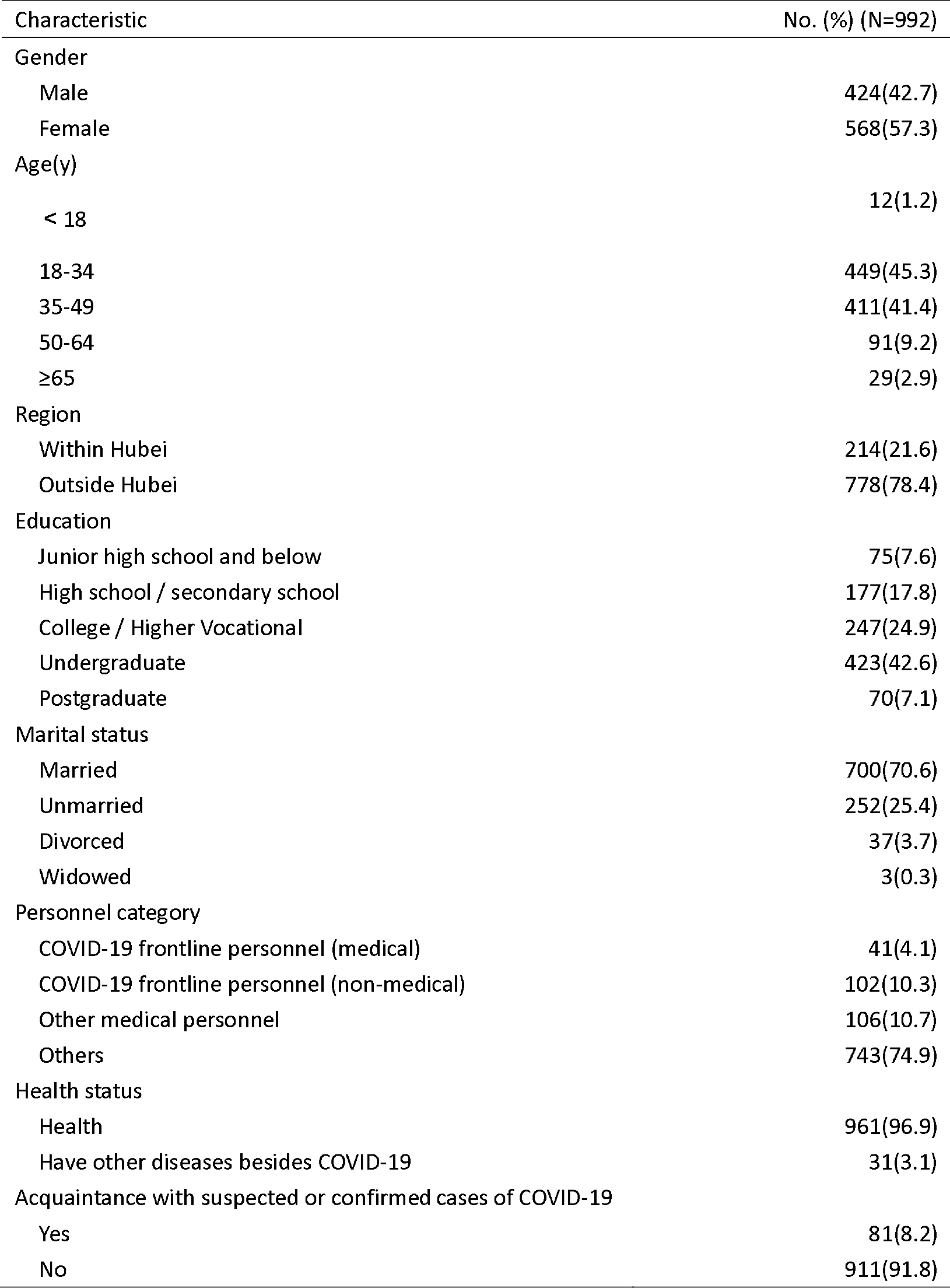
Characteristics of quarantined persons who responded to the survey

| Characteristic | No. (%) (N=992) |
| --- | --- |
| Gender |  |
| Male | 424(42.7) |
| Female | 568(57.3) |
| Age(y) |  |
| < 18 | 12(1.2) |
| 18-34 | 449(45.3) |
| 35-49 | 411(41.4) |
| 50-64 | 91(9.2) |
| ≥65 | 29(2.9) |
| Region |  |
| Within Hubei | 214(21.6) |
| Outside Hubei | 778(78.4) |
| Education |  |
| Junior high school and below | 75(7.6) |
| High school / secondary school | 177(17.8) |
| College / Higher Vocational | 247(24.9) |
| Undergraduate | 423(42.6) |
| Postgraduate | 70(7.1) |
| Marital status |  |
| Married | 700(70.6) |
| Unmarried | 252(25.4) |
| Divorced | 37(3.7) |
| Widowed | 3(0.3) |
| Personnel category |  |
| COVID-19 frontline personnel (medical) | 41(4.1) |
| COVID-19 frontline personnel (non-medical) | 102(10.3) |
| Other medical personnel | 106(10.7) |
| Others | 743(74.9) |
| Health status |  |
| Health | 961(96.9) |
| Have other diseases besides COVID-19 | 31(3.1) |
| Acquaintance with suspected or confirmed cases of COVID-19 |  |
| Yes | 81(8.2) |
| No | 911(91.8) |

### 2. Prevalence of anxiety symptoms by demographics of respondents

Table 2 summarized the prevalence of anxiety score according to demographics of respondents. According to clinical diagnosis standard in China and previous reports(Zung, 1986), 897 (90.42%) have normal SAS scores (<50), and 95 (9.58%) have elevated scores (≥50) indicating clinical significance of anxiety: 67(6.75%) are mild anxiety, 20(2.02%) are moderate anxiety and 8(0.81%) are severe anxiety. Anxiety scores in different age (F = 3.168, P = 0.013), education (F = 3.865, P = 0.004), health status (t = −3.043, P = 0.005), and personnel category (F = 5.802, P = 0.001) groups were statistically significant (P < 0.05). Immature adults are more anxious (mean = 46.33, median = 45.50, IQR 32-62) than other groups. Respondents with education lower than junior high school (mean = 41.11, median = 41, IQR 35-45) are more anxious than other groups. People with chronic diseases (mean = 44.06, median = 42, IQR 36-51) are more anxious. Frontline medical personnel are also more anxious (mean = 42.61, median = 41, IQR 34-44) which is matched with earlier report(Zhu et al., 2020).

**Table 2.**
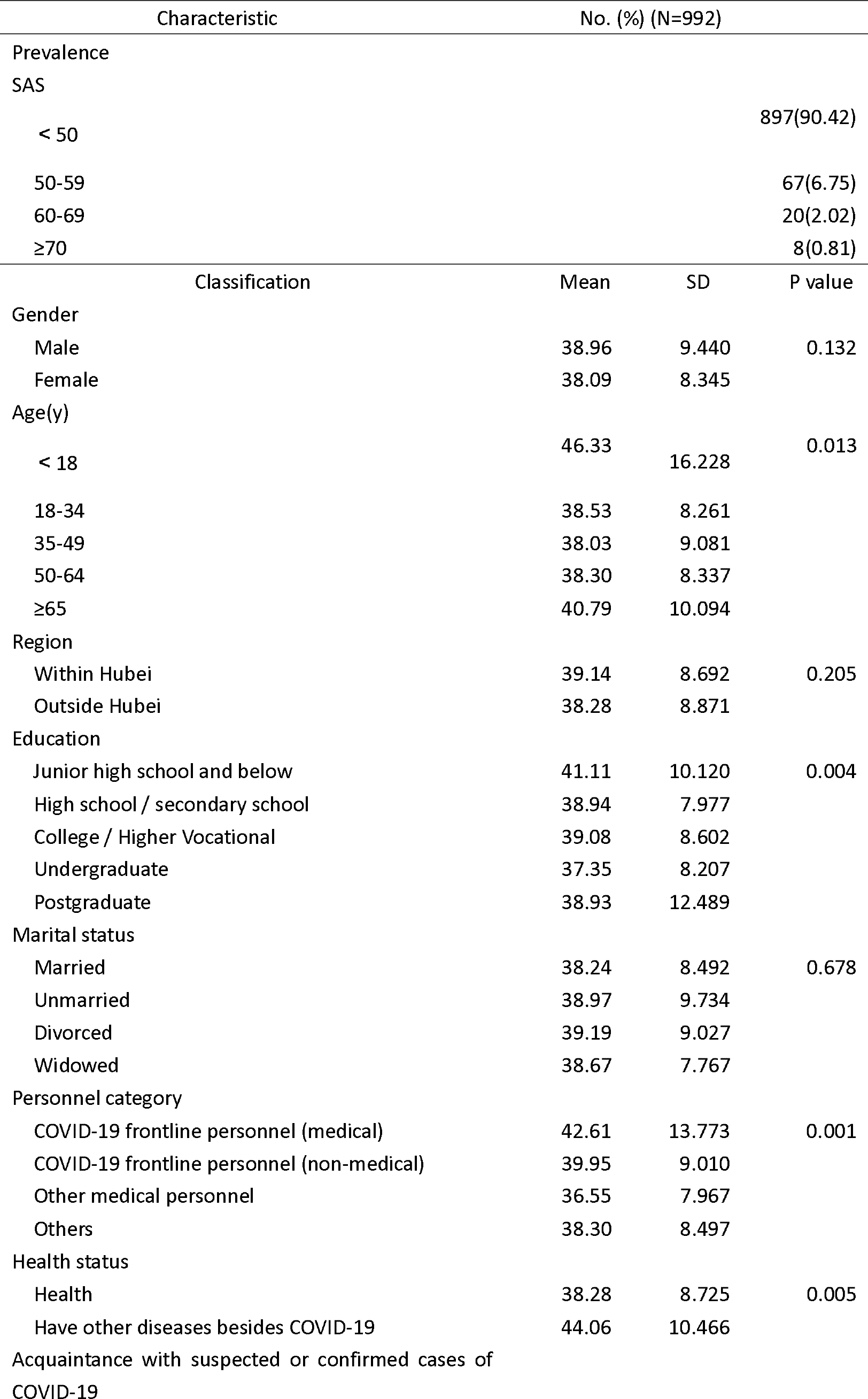
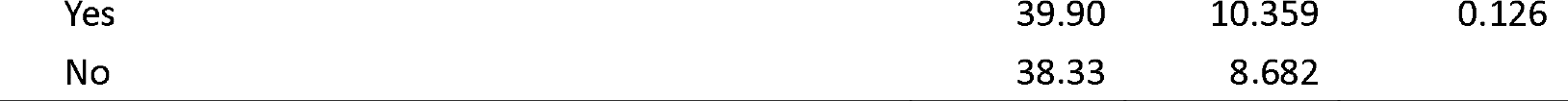
Prevalence of anxiety symptoms by demographics of respondents

Anxiety scores between different gender (t = 1.508, P = 0.132), region (t = 1.269, P = 0.205), marital status (F = 0.506, P = 0.678), and acquaintance with suspected or confirmed COVID-19 (t =- 1.531, P = 0.126) were not statistically significant among the respondents. The mild and not significant difference between respondents in and outside Hubei province was not expected because Hubei province has more than 80% infections in the whole country(National Health Commission of the People’s Republic of China, 2020b).

### 3. Different types of quarantine affect anxiety level differently

Multiple types of quarantine have been used during COVID-19 outbreak in China. Fig. 1 showed the proportion and SAS scores of respondents experienced different kinds of quarantines. Six kinds of quarantines were investigated in this survey: 1. Voluntary quarantine: stay at home voluntarily; 2. Semi-closed community: A permit is delivered to residents for limited number of entries and exits; 3. Fully-closed community: No one is allowed to enter or leave except for personnel responsible for supply dispensing; 4. Forced quarantine: Required to stay at home for certain periods; 5. Centralized quarantine: Quarantine at a designated place (e.g. a hotel); 6. Medical observation: Quarantine at a designated hospital. Because one person could experience multiple types of quarantines, his/her data might appear in different groups. Most of respondents (955/992, 96.27%) have at least one type of quarantine experience (Fig. 1A) and only 37/992 (3.73%) have no quarantine experience at all. Further analysis revealed that majority (29/37, 78.38%) of respondents without quarantine experience are frontline personnel for battling COVID-19, either medical or non-medical.

**Figure 1:**
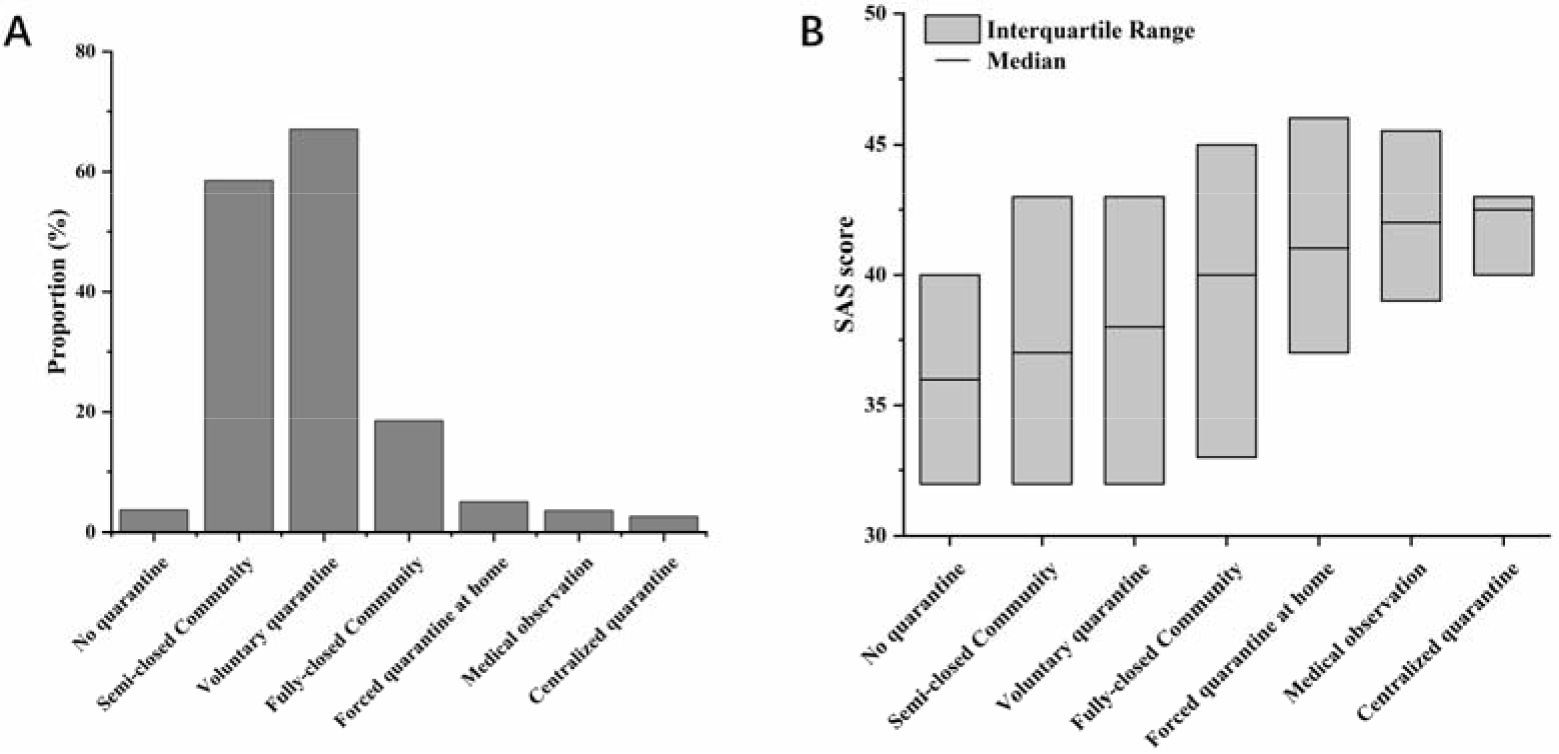
Proportion and anxiety scores of six groups of quarantine. A. Proportion of six types of quarantine. B. Box plot of anxiety scores in each group of quarantine. Interquartile range represents the range of 25%-75% scores.

Statistical results showed a significant difference (Fig. 1B, F = 5.132, p < 0.001) between different types of quarantine: no quarantine (median = 36, IQR 31.5 - 40), semi-closed Community (median = 37, IQR 32-43), voluntary quarantine (median = 38, IQR 32 - 43), fully-closed Community (median = 40, IQR 33 - 45), forced quarantine at home (median = 41, IQR 37 - 46), medical observation (median = 42, IQR 38.5 - 45.75), centralized quarantine (median = 42.5, IQR 39.25 - 43.5). This suggested that after quarantine, specific population might need medical assistance from professional psychologists.

### 4. Duration of quarantine does not significantly increase anxiety level

Previous studies suggested that longer duration of quarantine can result in worse psychological impacts(Brooks et al., 2020). Fig. 2 showed the proportion and SAS scores of different quarantine durations. Because the survey was taken in middle-late February and countrywide quarantine started in late January, most respondents (65.49%) have been quarantined for more than two weeks. Only 4.79% of respondents have quarantine duration less than 7 days. Statistical results revealed a mild but not significant difference between anxiety scores in groups with different durations (Fig.2B, F = 1.644,P = 0.178). The lowest score was found in group with less than 7 days (Mean = 36.87, Median = 36, IQR 32-42). But for duration longer than 7 days, all three groups have very similar scores: 8-14 days (mean = 38.14, median = 38, IQR 32-43), 15-28 days (mean = 39.04, median = 38, IQR 32-43), longer than 28 days (mean = 39.83, median = 38, IQR 33-43). This unexpected finding is encouraging but could be a result of many different reasons which we will discuss in details later.

**Figure 2:**
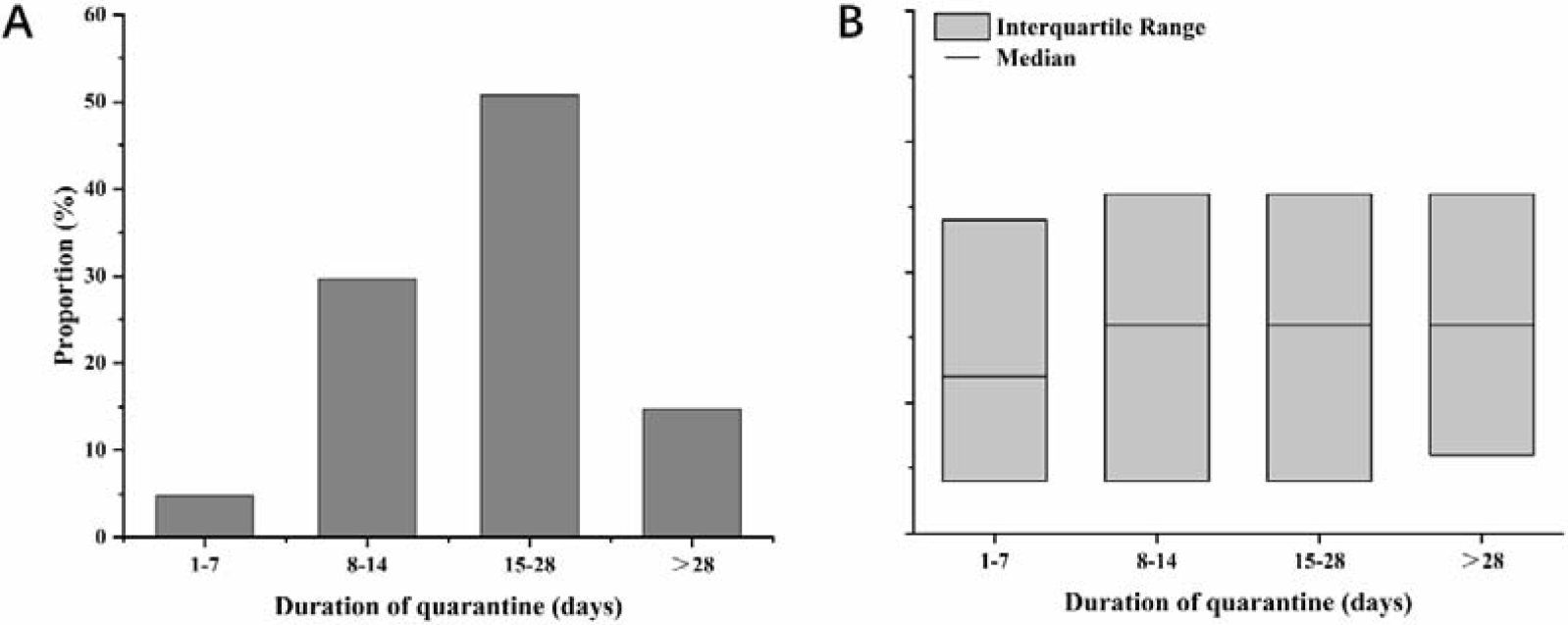
Proportion and anxiety scores of four groups of quarantine duration. A. Proportion of different durations of quarantine. B. Box plot of anxiety scores in each group of quarantine duration. Interquartile range represents the range of 25%-75% scores.

## Discussion

In the human history of fighting epidemics, there is very few successful cases that the outbreak was stopped by specific drugs or vaccines. This is usually caused by the rapid outbreak and the relatively slow development of specific drugs. When many companies finally developed SARS vaccines that could be used in clinical trials, the epidemic was almost over(Jiang et al., 2005). On one hand, we should keep developing effective drugs and therapies for COVID-19 as soon as possible; on the other hand, we should also consider using reasonable and effective quarantine methods (regional or national) to slow down the spread of the epidemic — buy some time for pharmaceutical research and production. Through this study, we found that countrywide quarantine in China does lead to increase of anxiety levels in certain population. For example, frontline personnel for battling COVID-19 (medical or non-medical) have higher risk of anxiety. These results are consistent with previous literature(Tam et al., 2004, Lee et al., 2018) and suggested more psychological assistance should be provided to specific populations. The more encouraging part of results is that compared with previous studies of quarantine(Jeong et al., 2016, Reynolds et al., 2008), the super large-scale quarantine in China did not lead to a significant change in anxiety level. The level of anxiety in Hubei Province, the most severely infected area in the world, was not significantly but only slightly higher than that outside Hubei Province (P = 0.205). Considering that Hubei province accounts for more than 80% of total infections in China and was the first quarantined province, these results are promising to the people in severed infected and strictly controlled areas such as Daegu in Republic of Korea or Lombardy in Italy.

However, the results we found here might not be easily replicated in other countries because there are many different reasons underlying the findings in study. (1). The survey was taken between February 19th to February 26th, a time window where the outbreak of COVID-19 has been largely controlled in China (Fig. S1). At this stage, the initial panic of COVID-19 outbreak has gone and many people in the country were still under quarantine. This ensured more accurate reflection of psychological influence induced by quarantine which is the main purpose of this study. But it is still unclear about what happened in the first a few days since outbreak. (2). The Chinese people and government worked extremely hard to overcome potential shortage of medical resources and food supplies. The financial aid provided by the government are also very helpful, including fully covered cost for diagnosis and treatment. These could be crucial important to reduce the risk of social panic. (3). Very different from previous epidemic outbreak like SARS in 2003, the whole world has become an information society in the past 20 years. Whether it is voluntary or forced quarantine, people are still well connected with the outside world through Internet and smart phones. (4). As we mentioned earlier, the effect of equity theory could also be helpful to ease the nervous and other negative emotions during quarantine.

The limitations of our study and implications are as follows. (1). Compared with super large-scale number of people affected by the countrywide quarantine, the sample size of our survey was very small. Thus, our results might not be able to fully reveal the true situation. (2). Although there is not much we can do, the form of online survey naturally ignored certain population, especially those aged adults who could be more vulnerable to COVID-19(Wu and McGoogan, 2020) but do not use smart phones. From our survey, only a very small portion of respondents (2.9%) are older than 65 years. (3). As we mentioned earlier, anxiety level could be a mixed result of many different factors. It is almost impossible to find out the pure contribution of quarantine to anxiety level. For example, worries about potential infection or financial problem could also elevate anxiety score(McAlonan et al., 2007, Zheng et al., 2005). Some respondents even leaved message to express their concerns about childcare and education due to school suspensions.

## Data Availability

The data used in this study are available on request from corresponding author.

## Acknowledgements

We sincerely appreciate all the people especially doctors and nurses all over the world who are heroically battling COVID-19 now. Dr. Yi Zhou received funding from Natural Science Foundation of China (31771152, 31970932). Juan Qiao received funding from Xuzhou Science and Technology Bureau of Jiangsu Province (KC18172).

## Data Availability Statement

The data used in this study are available on request from corresponding author.

## Ethical Statements

This study was conducted with informed consent of the respondent and has been approved by ethics committee of Xuzhou Oriental People’s Hospital. Questionnaires are collected anonymously to ensure that personal privacy is not disclosed. Secure encryption provided by “Questionnaire Star” was performed throughout the entire process including data collection, transmission, and release

## Conflict of interests

The authors declare that they have no conflict of interest.

---

**Figure S1.**
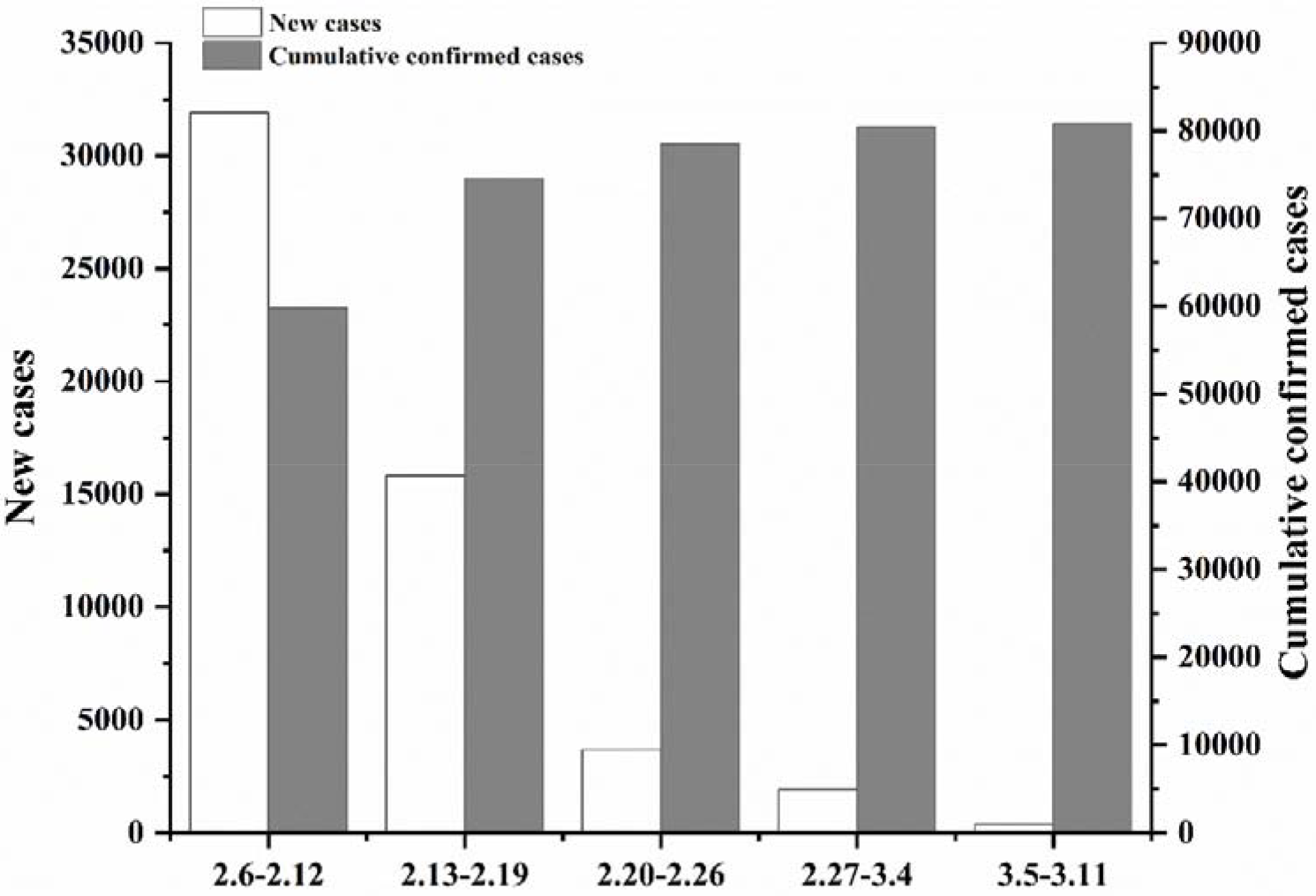
Epidemic situation before and after the survey (February 6^th^ – March 11^th^) The survey was performed between 12:00PM February 19^th^ 2020 and 12:00PM February 26^th^ 2020 (UTC+8). The number of cumulative cases was still increasing but the number of new cases was dropping rapidly when the survey is conducted. The numbers were collected from website of National Health Commission of PRC.

## Notes

### Competing Interest Statement

The authors have declared no competing interest.

